# Mothers of autistic children: a study of their experiences with child-protection social services, and allegations of fabricated or induced illness

**DOI:** 10.1101/2025.10.13.25337881

**Authors:** S. K. Crockford, A. L. Pohl, M. Blakemore, C. Allison, S. Baron-Cohen

**Author notes:** joint first authors (S. K. Crockford, A. L. Pohl). Author contact details: Corresponding author: Sarah K. Crockford Autism Research Centre University of Cambridge Department of Psychiatry, Douglas House, 18b Trumpington Road Cambridge CB2 8AH UK, **Email:**.

## Abstract

**Background**: Studies of autistic parenthood have consistently highlighted parents’ communication with health, education, and social services providers about their child as a source of stress due to stigmatization and discrimination. Our advisory panel suggested that mothers of autistic children might be at risk of unwarranted involvement with social services and unfounded accusations of Fabricated or Induced Illness (FII) i.e., fabricating illness in their child. The current study set out to explore these concerns. **Aims**: (1) To examine the degree of social service involvement in mothers who have an autistic child; (2) To quantify allegations of FII in mothers who have an autistic child; and (3) To test if either social services involvement or FII allegations varied as a function of whether the mother was herself autistic. **Methods**: N = 242 mothers of autistic children participated, comprising 3 groups: 101 autistic mothers with a formal diagnosis of autism; 67 mothers who self-identified as autistic; and 74 non-autistic mothers. All mothers completed a survey about involvement with social services and completed the short Autism Spectrum Quotient (AQ-10). All 3 groups were comparable on education, employment and marital status. **Results**: Over 20% of mothers of autistic children, regardless of maternal diagnosis, reported being investigated by social services for child welfare concerns. We also found that 2.48% (N=6 of N=242) of all mothers, regardless of diagnosis, reported an investigation of FII, and around 5% (N=12 of N=242) reported an allegation or investigation of FII. **Conclusions**: There is an alarmingly high frequency of social services involvement and allegations of FII in mothers of autistic children. Local authorities should consider having a specialist team conduct needs assessments for autistic children as parent-child dyads affected by autism are a large and growing population of social services users.

## Introduction

Advocates have long called for better support of autistic individuals across the lifespan by using social models of disability to adapt the world to neurodiverse people (1). However, we are only beginning to describe the needs of autistic adults (2). With autistic co-creators we identified a knowledge gap – autistic parenting; more specifically, motherhood – and co-designed a survey to report on the shared experiences of autistic mothers (3). As we and others have found, autistic parents’ communication with health, education, and social work practioners about their child is a source of stress: Autistic parents feel stigmatized and discriminated against and often end up in conflict with providers (3–5). During the survey design, our autistic co-creators raised the issue of discrimination in social services and care proceedings (please see the Supplementary Material for an explanation of the child welfare and protection system in the United Kingdom); others have since validated these reports in research samples (6,7). The purpose of this paper is to present exploratory data on autistic families’ involvement in care proceedings within the United Kingdom (UK) to generate discussion on the sensitive issues of justice and equity in care proceedings for parent-child dyads where one or both are affected by autism.

A brief word on the epidemiology of autism – and its influence on our understanding of autism – provides much-needed context for the fledgling literature on autistic parenting. Autism is more diagnosed in men, and women are underdiagnosed and diagnosed later in life than men (8,9). We have no national surveillance data on the number of adults that are autistic in any western country, but recent modelling suggests a prevalence of 2.2% in the United States, equating to nearly 5.5 million adults in the United States alone (10,11). As recently as 2015, only 2% of NIH research dollars in autism went towards adults (12). With no adult prevalence data and limited federal funding, public knowledge of and about autistic adults will remain biased towards antiquated, child-like stereotypes (12). Autism is also highly heritable and autistic traits exist on a continuous distribution in the general population; both diagnosable autism as well as high levels of autistic traits will aggregate in genetically-related families (13). In practice, this means that autistic parents or those with high levels of autistic traits will tend to raise autistic or high levels of autistic traits children, though some autistic parents raise non-autistic children, and some non-autistic parents raise autistic children.

Qualitative studies have revealed that parenthood is a desirable, fulfilling, and joyful experience for most autistic parents interviewed (4,5,14,15). Nuanced experiences specific to autistic parents are beginning to be described, including (i) that autistic parents have special insight into the needs of their autistic children through shared lived experiences, (ii) that autistic families and homes are a cocoon from the outside world, and will have different prioritization of needs and values based on the lived experience of autism, and (iii) that interactions with health, education, and social professionals regarding one’s children are stressful and that the autistic parent must fiercely advocate for their child’s needs in these scenarios (4–6,14,15). The struggle to interact with professionals – which Marriott *et al.* concisely coined “*the struggle to be heard, believed, and supported”*(*5*) – is the domain of autistic parenthood most urgently in need of research, as misalignment of parent-professional beliefs can have negative outcomes including missed diagnoses, missed opportunities for support or intervention, and the placement of children in state custody. Autistic parents are more likely to have autistic children who require additional support, and as one route to supportive services in the UK is through a parent-, healthcare professional-, or educator-initiated *needs assessment* conducted by social services, it follows that autistic children and autistic parents are coming into contact with social services (which has responsibility for child protection) more frequently than their non-autistic counterparts in the UK. Although though specific rates of needs assessments among autistic vs non-autistic children in the UK are not known, as of 2024 13.6% of children assessed as ‘in need’ in the UK were recorded as having a disability, of which 44.2% were recorded as being autistic (16).

Conversations with professionals about one’s child are an unavoidable part of parenthood, and present the autistic parent with a dilemma: do I disclose my diagnosis of autism in this conversation? Parents who choose to disclose may be rebuked or disbelieved (15); alternately, disclosure can lead to judgement and stigmatization (15). This experience is pervasive: 60% of our sample of 350 formally diagnosed autistic mothers had their diagnosis questioned in conversations with professionals about their child, and greater than 80% of autistic mothers worried that professionals’ attitudes would change if a diagnosis was disclosed (3). The alternative is to not disclose a diagnosis, removing the possibility that parental behaviours could be properly contextualized through a diagnosis of autism, and therefore subjecting autistic parents to non-autistic standards of parenting (7). As autistic parents reflect upon their own childhood experiences (*e.g.* being bullied at school, experiencing sensory overload at school), they may reject some normative parenting standards (*e.g.* children attending mainstream school) to protect an autistic child from a distressing situation – in one mother’s case, the decision to pull her child from mainstream school landed her on a child protection plan (4,7).

Most direly, a parent’s disclosure of autism could lead to increased scrutiny and false allegations of fabricated or induced illness (FII), also referred to as Munchausen Syndrome by Proxy (MSP) or Factitious Disorder Imposed on Another (FDIoA). In their review of FII in the UK, Gullon-Scott and Long (17) describe how a single article incorrectly conflated parental autism with FII (18) by misinterpreting the results of a 3-case series (19). This claim was parroted without fact-checking (20), and has subsequently been enshrined in best-practice policy guidelines in the UK (21). FII often leads to the placement of the child in care (22); therefore, FII misdiagnosis could violate the human right of autistic parents to have a family without unwarranted intervention from the state.

We do not know what percent of autistic children are evaluated by social services for protection concerns, put on a child protection registry, or investigated for being the victim of FII in the UK, nor do we know if maternal autism diagnosis influences these rates. Here we report preliminary data on these questions in the UK which we obtained from a survey sent to mothers of autistic children. Unfortunately obtaining unbiased complementary data on non-autistic children and their parents would require telephone-based sampling, which was not reasonably achievable in this study. We predicted that a high number of mothers of autistic children would report being social services involvement, having their child forcibly removed by the state, and being accused of FII compared to epidemiologic data from the general population. We predicted these risks would be greater in autistic compared to non-autistic mothers.

## Methodology

We developed an online survey that concerned various aspects of motherhood, using a Public and Patient Involvement and Engagement (PPIE) model, which we have detailed elsewhere (3). Briefly, an advisory panel of six autistic mothers met with the research team four times to help design, disseminate, and interpret the study. All panel members were reimbursed for their time and travel according to best practices.

We embedded questions regarding social services in a larger survey but are separately reportingly the results here as (i) these questions were only shown to participants from the UK, and (ii) we felt that discrimination in care proceedings was a sensitive issue that merited its own publication for discussion. Items included questions about whether participants ever had their child investigated for risk of harm, have ever been called into a meeting with social services, ever had their child placed on a child protection register, and whether a child of theirs has even been placed in foster care or forcibly adopted, and whether they have ever been investigated for FII. Questions included in this section of the survey were a combination of forced choice answers, requiring a yes/no or agree/disagree statement, and free text. The full list of survey items is included in the Supplementary Materials.

We made the survey available online and distributed it via autism support groups and the Cambridge Autism Research Database (CARD). We recruited mothers of autistic children and separated the mothers by diagnosis for analysis. In addition to the UK-specific rationale discussed in the introduction, more autistic than typically developing children receive child protective services in the United States (US) (23,24). Therefore, to examine differences based on maternal autistic diagnosis, the child’s diagnosis must be held constant, as an autistic child and a non-autistic child do not have the same chance of having a needs assessment by social services in the UK, nor do they have the same chance of being found at-risk of harm, based on the US data. 258 non-autistic mothers and 410 autistic (diagnosed and self-identified) mothers completed the survey in full. We presented questions regarding interactions with social services only to participants based in the UK, using the geographic localisation of the participant’s IP address. Our final sample was reduced to 101 clinically diagnosed autistic mothers, 67 mothers who self-identified as being autistic, and 74 non-autistic mothers. Diagnostic information was obtained by self-report, and we validated groups through the AQ-10, which measures autistic traits and is adapted from the full version of the Autism Spectrum Quotient (25,26). The cut-point for possible autism is 6 or above, with a higher score representing more autistic traits. Participants were not reimbursed for their participation in the survey to discourage falsified responses. Given the high prevalence of sex/gender discordance in autism (27), we encouraged participants to self-report sex and gender, and did not restrict participation by sex or gender identity, as both Female-to-Male (FTM) and Male-to-Female (MTF) transgender individuals could identify with the label of “mother”, depending on whether one’s definition of being a mother is linked to sex assigned at birth or gender respectively.

Furthermore, it is illegal in England to disclose information regarding legal care proceedings; therefore, we obtained clearance to proceed with this research from Her Majesty’s Court and Tribunal Services in addition to ethical approval from the Psychology Research Ethics Committee at the University of Cambridge. Data from the survey were analysed using R Studio. Differences were calculated with one-way ANOVA, paired t-tests, and Pearson’s *X*^2^ test, with a simulated p-value based on 2000 replicates, where appropriate. Deductive coding was used to classify the reason(s) for child protection registration, fostering, and adoption orders and two authors (SKC and AP) agreed on all codes.

## Results

### Group demographics

Non-Autistic, self-reported diagnosed, and self-identified autistic mothers were demographically similar (see Table 1). Only current maternal age was significantly different between groups, with non-autistic mothers being older than autistic and self-identified autistic mothers. However, on average, all groups of mothers in the study were about forty-years of age. All self-identified autistic mothers reported being biologically related to their children, while 1% of autistic and 4% of non-autistic mothers were not biological mothers of at least one child.

**Table 1.** Demographic characteristics.

|  | **Diagnosed**<br>**Autistic** | **Self-**<br>**Identified**<br>**Autistic** | **Non-**<br>**Autistic** | ***p*-**<br>**value** |
| --- | --- | --- | --- | --- |
| Age (years) | 43.4 ± 7.8<br>[27- 68] | 43.3 ± 7.0<br>[24 – 59] | 46.3 ± 9.06<br>[25 -74] | **0.037** |
| Sex (%) |  |  |  | 0.745 |
| Female | 100 (99%) | 66 (99%) | 74<br>(100%) |  |
| Male | 1 (1%) | 1 (1%) | - |  |
| Gender (%) |  |  |  | 0.190 |
| Female | 96 (95%) | 64 (95%) | 71 (96%) |  |
| Male | 2 (2%) | - | 3 (4%) |  |
| Other | 3 (3%) | 3 (5%) | - |  |
| Education |  |  |  | 0.116 |
| Some secondary / high school | 8 (8%) | 3 (5%) | 4 (5%) |  |
| Completed secondary / high school | 17 (17%) | 17 (25%) | 21 (28%) |  |
| Completed some of an undergraduate degree | 21 (21%) | 13 (19%) | 4 (5%) |  |
| Completed an undergraduate degree | 25 (25%) | 18 (27%) | 18 (24%) |  |
| Completed a postgraduate / graduate degree | 30 (28%) | 16 (24%) | 27 (36%) |  |
| Marital Status |  |  |  | 0.283 |
| Single | 13 (13%) | 8 (12%) | 7 (9%) |  |
| Married | 53 (53%) | 37 (55%) | 46 (62%) |  |
| Civil partnership | 1 (<1%) | - | - |  |
| Divorced | 13 (13%) | 9 (13%) | 3 (4%) |  |
| Widowed | 2 (1%) | 2 (3 %) | 1 (1%) |  |
| Separated | 7 (7%) | 7 (11 %) | 4 (5%) |  |
| Long-term partner | 13 (13%) | 4 (6 %) | 13 (18%) |  |
| Other | 1 (<1%) | - | - |  |
| Living with current partner (if applicable) |  |  |  | 0.592 |
| Yes | 68 (100%) | 41 (100%) | 57 (98%) |  |
| No | - |  | 1 (2%) |  |
| Does partner have autism? (if applicable) |  |  |  | 0.174 |
| Diagnosed | 7 (10%) | 1 (2%) | 1 (2%) |  |
| Suspected | 25 (37%) | 14 (34%) | 18 (31%) |  |
| No | 36 (53%) | 26 (63%) | 39 (67%) |  |
| Single Parenthood |  |  |  | 0.072 |
| Currently a single parent | 28 (28%) | 24 (36%) | 13 (18%) |  |
| Have been a single parent in the past | 19 (19%) | 15 (22%) | 13 (18%) |  |
| Never have been a single parent | 54 (53%) | 28 (42%) | 48 (65%) |  |
| Age at first birth (years) | 27.6 ± 6.3<br>[15 – 41] | 28.2 ± 6.3<br>[16 – 42] | 29.7 ± 6.6<br>[18 – 45] | 0.098 |
| Number of children | 2.5 ± 1.9<br>[1-12] | 2.5 ± 1.3<br>[1-6] | 2.3 ± 1.1<br>[1-6] | 0.551 |

*Numeric variables are given in the form mean ± standard deviation; [range].*

### Distribution of Autistic traits

We compared scores on the 10-item version of the Autism Spectrum Quotient (AQ-10) to validate participants’ self-reported diagnosis. There was a significant effect of diagnostic group on AQ10 scores (F(2) = 193.7, p <2e-16), with diagnosed autistic mothers scoring highest (mean = 7.65, sd = 1.38, [4 – 10]), followed by self-identifying (mean = 6.56, sd = 1.78, [3 – 9]), then non-autistic mothers (mean = 2.92, sd = 1.64, [1 – 8]). A follow up Tukey comparison of means showed significant differences (p < 0.01) between each of the three groups.

### Interacting with social services

Over 20% of mothers of an autistic child, regardless of maternal diagnosis, reported having had at least one child assessed by social services for being at risk of harm. There was no statistical difference between the groups (χ^2^ = 0.39, p =0.850) (Table 2).

**Table 2.**
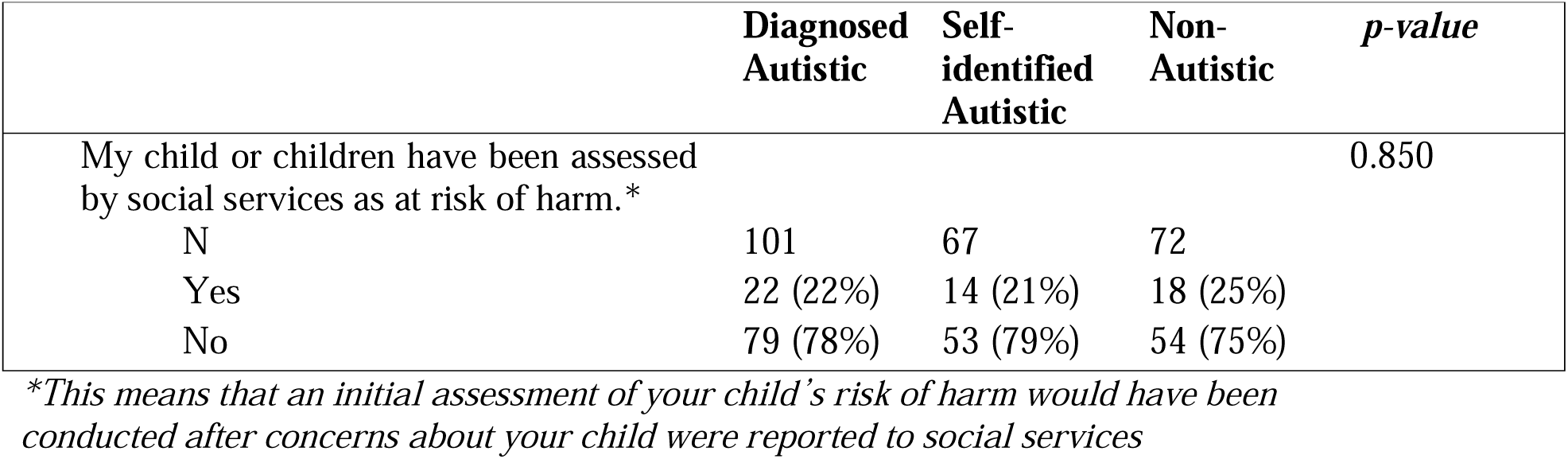
Number of mothers who had a child assessed by child services for at-risk of harm.

There was also no statistical difference between groups in the proportion of mothers called into a meeting with social services (χ^2^ = 0.58, p-value = 0.764). We also asked these mothers whether they fully understood the purpose of the meeting and its legal implications. A majority of mothers across all groups did not fully understand the implications or legal ramifications of their meeting with social services (χ^2^ = 1.93, p-value = 0.375) (Table 3). However, it is important to note that the subsample of mothers across all three groups who responded to this question is quite small, N = 21 for autistic mothers, N = 17 for self-identified autistic mothers, and N = 15 for non-autistic mothers.

**Table 3.** Number of mothers called into a meeting and who understood their meeting with social services.

|  | **Diagnosed**<br>**Autistic** | **Self-**<br>**identified**<br>**Autistic** | **Non-**<br>**Autistic** | *p-value* |
| --- | --- | --- | --- | --- |
| I have been called to a meeting with social services about my child. |  |  |  | 0.764 |
| N | 101 | 67 | 72 |  |
| Yes | 21 (21%) | 17 (25%) | 15 (21%) |  |
| No | 80 (79%) | 50 (75%) | 57 (79%) |  |
| When I was called to the meeting with social services, I didn't understand what the purpose of the meeting was or what the legal implications of the meeting were. |  |  |  | 0.375 |
| N | 21 | 17 | 15 |  |
| Agree | 15 (71%) | 9 (53%) | 11 (73%) |  |
| Disagree | 6 (29%) | 8 (47%) | 4 (27%) |  |

### Registration and removal of children

We found no significant differences between diagnostic groups on the number of mothers who reported having their child placed on the Child Protection Register (χ^2^ = 1.73, *p* = 0.402), under a foster order (χ^2^ = 1.44, *p* = 0.557) or adoption order (χ^2^ = 1.38, *p* = 0.563). Raw totals and percentages of mothers in each group are reported in Table 4. Participants also provided free-text explanations for why their child was placed on the register, fostered, or adopted. Of 23 total observations, emotional abuse was given as a reason for child registration in N = 9 cases, followed by neglect (N = 7), a child’s behavioural issues putting themselves or their siblings at risk (N = 4), domestic violence perpetrated against the mother (N = 3), FII (N = 2), physical abuse (N = 1) and child abuse not further specified (N = 1). Three participants did not provide a reason for their child’s placement on the register. Overall, a similar pattern of responses was given for children being fostered or adopted, with allegations of neglect and emotional abuse dominating but with domestic abuse and the child’s behavioural issues also represented.

**Table 4.** Number of mothers who reported having a child placed on a register, a foster order or an adoption order.

|  | **Diagnosed**<br>**Autistic** | **Self-**<br>**identified**<br>**Autistic** | **Non-**<br>**Autistic** | ***p-value*** |
| --- | --- | --- | --- | --- |
| Child Register |  |  |  | 0.402 |
| N | 101 | 67 | 70 |  |
| No | 94 (93%) | 60 (90%) | 61 (87%) |  |
| Yes | 7 (7%) | 7 (10%) | 9 (13%) |  |
| Foster Order |  |  |  | 0.557 |
| N | 100 | 67 | 71 |  |
| No | 96 (96%) | 65 (97%) | 66 (93%) |  |
| Yes | 4 (4%) | 2 (3%) | 5 (7%) |  |
| Adoption Order |  |  |  | 0.563 |
| N | 101 | 67 | 72 |  |
| No | 96 (95%) | 66 (99%) | 69 (96%) |  |
| Yes | 5 (5%) | 1 (1%) | 3 (4%) |  |

### False allegations by social services

We also asked mothers whether allegations made towards them by social services were untrue or inaccurate. Across the three groups, 22% (N=22) of diagnosed autistic mothers found this to be the case compared to 22% (N=15) of self-identified autistic mothers and 19% (N=14) of non-autistic mothers. When we compared the groups, no statistical difference emerged (χ^2^ = 0.22, *p* = 0.906).

### Allegations of fabricated illness

We found 6 out of 242 (2.48%) mothers had been investigated for FII. There were no differences in rates of investigations of FII in the three groups: 1% (N=1) of diagnosed autistic mothers, 3% (N=2) of self-identified mothers and 4% (N=3) of non-autistic mothers had been investigated for FII (χ^2^ = 1.80, *p* = 0.456). However, when we reviewed free text responses to the question *“Have social services ever said things about you that you felt were untrue or inaccurate?”* an additional six participants reported allegations or rumours of FII that were never formally investigated. The following statements were reported by these six additional participants: *“they were potentially heading towards a MSBP/FII case, they were accusing me in the professional network of inventing or exaggerating my children’s school difficulties”, “making him Autistic as reflecting own problems/autism on to him”, “She’s making up his autism”, “my child didn’t have additional need[s]…I over medicated my child and then accused of under-medicating my child”, “heard via the school nurse that the team … were considering fabricated illness being involved”, and “accused by a newly trained psychologist of [causing] my 3^rd^ child’s anxiety.”* As a sensitivity analysis, we analysed allegations of fabricated illness re-classifying these six individuals – there were no differences in the three groups: 3% (N=3) of diagnosed autistic mothers, 4% (N=3) of self-identified autistic mothers and 8% (N=6) of non-autistic mothers reported allegations or investigations of FII (χ^2^ = 2.55, *p* = 0.28).

## Discussion

In 2016, around the time this study was conducted, 6% of children and young people met the criterion of being ‘in need’ in the UK, meaning they would qualify for a *needs assessment* by social services (28). By legal definition, all children with a disability, including autism, are ‘in need’ (28). In 2015, when our data were collected, autistic children accounted for 30.5% of all children in need with a reported disability in the UK. By 2024, the latest currently available reporting from the UK’s Office of National Statistics, the proportion of autistic children among children in need with a reported disability increased to 44.2% (16). Autistic children and their parents are thus clearly a large and growing fraction of social services users. In our sample, 1 in 5 mothers of autistic children report being investigated by social services for placing their children at-risk of harm. Of these 1 in 5 mothers, 43% will have their child placed on a child protection register (N = 23 total mothers who reported being placed on a child protection register of N = 54 total mothers who reported being investigated by social services). Population prevalence estimates in England in 2015 for children defined as being in need and children placed on child protection plans were 338.2/10,000 (3.4%) children and 43.1/10,000 (0.4%) children respectively (16). Thus, our results suggest a significant increase in inquiries and registrations in families with autistic children, regardless of maternal diagnosis, compared to the general population.

Autistic people are vulnerable to abuse and crime (29). The vulnerability of autistic children to abuse, neglect and bodily harm has unfortunately been recognized via high-profile familicides of autistic children, including most notably Katherine “Katie” McCarron, a toddler who was suffocated in 2006 by her mother. Autistic children are overrepresented in child protection services and foster care in the US, with neglect being less frequent and physical abuse being more frequent than non-autistic children under protection (23,24). However, in a Medicaid sample of 40 million disadvantaged children receiving state-funded healthcare, there was only a 3-fold difference between the rate of autistic (8.1%) and non-autistic (2.6%) disadvantaged children in social care (24). The history of intense public scrutiny of the social workers involved missed abuse cases in the UK – see “Baby P’ – has led to an intense fear of missing an abuse case among child protection social workers in the UK, in addition to other pressures they face such as understaffing and threats of personal violence (30). By comparing international rates of autistic and non-autistic children on child protection plans or in care, it may be possible to see whether autistic children in the UK are entering protection at disproportionately high rates. If so, this could suggest that discrimination against parents of Autistic children is contributing to the increase beyond the elevated baseline risk expected for autism.

Parents with an intellectual disability are at increased risk of scrutiny by social services, demonstrating that discrimination exists in care proceedings, and we hypothesized that autistic mothers would be more likely to have a child in protection than non-autistic mothers (31). We found no statistical differences between our groups at any stage of the child protection process. As both autistic parents and non-autistic parents of Autistic children feel disbelieved when interacting with professionals (4–7, 32), the struggle to be believed by professionals – and potential downstream consequences in care proceedings – may be shared by all parents of autistic children. Further work could contrast the experiences of parents of children with autism to those with a more visible disability, such as cerebral palsy.

An alarming number of mothers reported allegations of FII. The reported incidence of FII in the UK and the Republic of Ireland was 0.5/100,000 in children under 16 years of age and 2.8/100,000 in children under the age of 1 in 1996 (33). In 2006, the prevalence was estimated at 30 in 100,000 in Germany in a tertiary health care center (34), and 2 in 100,000 in New Zealand in 2001 (35). By comparison, a conservative 2.5 in 100 mothers of autistic children reported being investigated for FII, and six other participants mentioned rumours of FII, which would bring our estimate to 5 in 100. We found that allegations of FII are *two to three orders of magnitude higher in mothers of Autistic children in the UK* than estimates of FII occurrence in the general population. Substantiated cases of FII in national surveillance in New Zealand most commonly included seizures and apnoea as the primary presentation, and autism was noticeably absent from the list of presenting symptoms (35). Further research is needed to understand the number of and reasons for FII investigations in autistic children.

## Limitations

This study was limited by only including mothers with at least one autistic child; we did not actively recruit mothers of typically developing children due to practical considerations and the differences in baseline vulnerability between autistic and typically developing children. Second, the questions in the survey were exploratory and therefore we did not enquire in detail about social service involvement. Further research should examine individual cases in more detail to understand the nuances of interactions between autistic families and social services. A final limitation of this study is that we did not seek the input of child protection social workers who offer a key perspective on this issue. Safeguarding children is the priority; however, proceedings must be free of bias and accessible to parents with disabilities to ensure the best outcomes for children. We hope that these findings will open a discussion and future research on how to improve child protection assessments in families with an autistic parent or child and open a dialogue between those practicing in child protection and the autistic community.

## Conclusion

Working with autistic mothers, we identified that 1 in 5 mothers of autistic children in the UK are investigated by social services for child welfare concerns. We found FII allegations against mothers of autistic children were much higher than the incidence of FII in the general population. There was no variation by maternal diagnosis in any stage of child protection investigation. We call for international data on the prevalence of autistic children under protection so that governments with outlying rates of autistic children in protection can investigate their practices for bias. Child protection is a pervasive yet woefully understudied topic in autism, and we look forward to exchanging ideas with physicians, psychologists, health visitors, nurses, social workers, and educators working in child protection.

## Declarations

### Ethics approval and consent to participate

Ethical approval for this study was granted by the Psychology Research Ethics Committee, University of Cambridge (Reference number: PRE.2015.049). Additional ethical approval for the study was also granted by Her Majesty’s Court and Tribunal Services (HMCTS). Participants gave their informed consent to take part in this study by agreeing to the terms of the study prior to accessing the online survey.

## Consent for publication

Not applicable.

## Availability of data and materials

The datasets generated during and/or analysed during the current study are not publicly available due to the sensitive nature of some of the data but are available from the corresponding author on reasonable request.

## Authors Contributions

SKC wrote the final manuscript, analysed and interpreted the data, contributed to the survey design, identified the need for an advisory panel during the study design, and helped with the implementation of the advisory panel. AP analysed and interpreted the data, developed and disseminated the online survey, secured the funding grant to cost the advisory panel, chaired the advisory panel, and co-wrote with SKC the final manuscript. MB developed the research focus of this study, aided in the dissemination of the survey and of the study findings and contributed to writing the manuscript. SBC and CA supervised the research, co-developed the survey and were major contributors to the writing of the manuscript. All authors read and approved the final manuscript.

## Acknowledgements

We acknowledge the contributions made by the members of our advisory panel, without whom this study would not have been possible. We are grateful to Rebecca Kenny, Rosie Holt, and Amber Ruigrok for helpful discussions.

## Supplementary material

### Experiences with Social Services

#### This section is only displayed to individuals accessing the questionnaire from a United Kingdom IP address

Have any of the following experiences happened to you? Please answer yes or no.

➢ My child or children have been assessed by social services as at risk of harm. This means that an initial assessment of your child’s risk of harm would have been conducted after concerns about your child were reported to social services. YES NO
➢ I have been called to a meeting with social services about my child. YES NO
  ○ IF ‘Yes’ is selected When I was called to the meeting with social services, I didn’t understand what the purpose of the meeting was or what the legal implications of the meeting were. Strongly Agree Agree Disagree Strongly Disagree
➢ My child or children have been placed on the child protection register. YES NO
  ○ IF ‘Yes’ is selected What was the reason for them being placed on the child protection register?
➢ My child or children have been temporarily taken out of my care under a fostering order. This means that your child or children would have been temporarily been placed in care. YES NO
  ○ IF ‘Yes’ is selected What was the reason for them being temporarily taken out of your care?
➢ I have been investigated for Munchausen Syndrome by Proxy or Fabricating Illness in my child or children. YES NO
➢ Have social services ever said things about you that you felt were untrue or inaccurate? YES NO
  ○ IF ‘Yes’ is selected What was said that you felt was untrue or inaccurate?

## Model of UK social services (at the time of this study)

The aim of this study was to gather information on whether autistic mothers were more likely than their non-autistic counterparts to be assessed, investigated, and have their children removed by social services. Every state-funded authority has its own regulations with regards to how it is allowed to intervene in childcare, with the focus of this study being on UK social services. Although the system in place may vary between England, Wales, Scotland and Northern Ireland, we would expect that on average, mother’s experiences will be relatively comparable.

According to guidelines published by the National Health Services (NHS, https://www.nhs.uk/conditions/social-care-and-support/services-for-children-and-young-people/) and the National Society for the Prevention of Cruelty to Children (NSPCC, https://www.nspcc.org.uk/), in line with the *Children Act of 1989* (CA, 1989), families undergo an initial assessment by social services. This can happen because either a parent and/or primary caregiver has requested a needs assessment in order to access additional support for their child or because someone outside of the family has reported the child as being at risk of harm. If the parents/primary caregivers do not consent to the assessment, a child assessment order can be issued by the LA (Local Authority).

Once the assessment has occurred, there can be a number of different outcomes: The child is deemed not to be at risk or in need of further support and is discharged from social services; the child is designated as a child ‘in need’, meaning that the child has significant health issues and/or is disabled and therefore the family requires further support; and/or there are serious concerns about the child’s safety and a strategy discussion takes place. The strategy discussion is a time during which professionals assess whether the child is at risk of harm and social services further investigate the family involved. This can lead to a *Section 47* enquiry where a decision will be made as to whether the child is at risk for significant harm. If this is the outcome, then a case conference will be held at which the parents and/or primary caregivers may be asked to attend and will include a variety of professionals involved to some extent with the family, i.e. clinician, social workers. The case conference will lay out a child protection plan, where attempts will be made to improve family life so that the child does not need to be removed from the parents.

If, following a number of case conferences, it is decided that child will be removed from their current parental and/or primary care giver’s care then the child’s carer will be invited to a pre-proceedings meeting. This is the final step before going to court and involves parents and/or primary care givers, the LA and lawyers. An effort is made to establish a formal agreement between parents and the LA. If no agreement can be found or parents do not follow the obligations laid out by the formal agreement, then the LA will pursue the family in court. Depending on the complexity of the case, this will either be in Family or High Court. The outcome of the court case will decide whether the child should be placed in temporary foster care, a permanent foster order, a placement order (where the child is placed somewhere temporarily whilst awaiting adoption) or an adoption order. These orders are court mandated and can be enacted without the consent of parents.

